# Postoperative Acute Kidney Injury is Associated with Persistent Renal Dysfunction: A Multicenter Propensity Matched Cohort Study

**DOI:** 10.1101/2024.06.06.24308455

**Authors:** Blaine Stannard, Richard H. Epstein, Eilon Gabel, Girish N. Nadkarni, Yuxia Ouyang, Hung-Mo Lin, Valiollah Salari, Ira S. Hofer

## Abstract

**Background:** The risk of developing a persistent reduction in renal function after postoperative acute kidney injury (pAKI) is not well-established.

**Objective:** Perform a multi-center retrospective propensity matched study evaluating whether patients that develop pAKI have a greater decline in long-term renal function than patients that did not develop postoperative AKI.

**Design:** Multi-center retrospective propensity matched study.

**Setting:** Anesthesia data warehouses at three tertiary care hospitals were queried.

**Patients:** Adult patients undergoing surgery with available preoperative and postoperative creatinine results and without baseline hemodialysis requirements.

**Measurements:** The primary outcome was a decline in follow-up glomerular filtration rate (GFR) of 40% relative to baseline, based on follow-up outpatient visits from 0-36 months after hospital discharge. A propensity score matched sample was used in Kaplan-Meier analysis and in a piecewise Cox model to compare time to first 40% decline in GFR for patients with and without pAKI.

**Results:** A total of 95,208 patients were included. The rate of pAKI ranged from 9.9% to 13.7%. In the piecewise Cox model, pAKI significantly increased the hazard of a 40% decline in GFR. The common effect hazard ratio was 13.35 (95% CI: 10.79 to 16.51, p<0.001) for 0-6 months, 7.07 (5.52 to 9.05, p<0.001) for 6-12 months, 6.02 (4.69 to 7.74, p<0.001) for 12-24 months, and 4.32 (2.65 to 7.05, p<0.001) for 24-36 months.

**Limitations:** Retrospective; Patients undergoing ambulatory surgery without postoperative lab tests drawn before discharge were not captured; certain variables like postoperative urine output were not reliably available.

**Conclusion:** Postoperative AKI significantly increases the risk of a 40% decline in GFR up to 36 months after the index surgery across three institutions.

## Introduction

Acute kidney injury (AKI) is a known complication of surgery, through mechanisms including hypoperfusion and direct tubular injury (1). Postoperative AKI (pAKI) has been associated with substantial consequences such as impaired cardiac function, pulmonary complications, volume overload, higher rates of systemic and wound infection, and delirium (1–6). Overall, pAKI has been associated with a 3 to 5-fold increased risk of postoperative mortality (7,8), longer length of stay, and increased rates of readmission (8,9).

Despite the many studies of the short-term complications of AKI, little work has been done regarding progression to long term renal dysfunction. In non-surgical patients, significant associations between AKI and new-onset or progression of existing CKD have been observed in the inpatient and intensive care unit (ICU) settings (10–12). However, the pathophysiology of pAKI may differ from causative factors in medical patients (e.g., sepsis), and thus the risk of developing a persistent reduction in renal function after pAKI cannot be directly extrapolated from non-surgical populations. The studies that did look at the progression of pAKI to CKD have been smaller single center studies, thus limiting generalizability (13,14). While AKI itself is associated with significant short term morbidity and mortality, if it is also associated with an increased incidence of CKD, that would substantially increase its long-term effects on patients.

In order to better understand the association between pAKI and progression to CKD we performed a multi-center retrospective propensity matched examination of the association of pAKI and longer term CKD. The primary outcome was a decline in glomerular filtration rate (GFR) of at least 40% (relative to preoperative baseline) (15), as measured at outpatient follow-up visits up to 36 months after discharge.

## Methods

Data were collected from departmental anesthesia data warehouses at Mount Sinai Hospital (MSH), the University of Miami (UM), and the University of California Los Angeles (UCLA). Institutional Review Board approval was obtained at each respective institution with waiver of informed consent (MSH: 19-01166-CR003; UM: 15-000518). All three institutions use a perioperative data warehouse from Extrico Health which has been described previously (16).

### Inclusion and Exclusion Criteria

Patients were included if they were ≥18 years old and underwent surgery at one of the three sites. Data were collected between years 2016-2021 at MSH, 2020-2024 at UCLA, and 2017-2023 at UM. No surgical specialties were excluded in the data extraction. Patients were included regardless of baseline CKD stage, which was controlled for in the analysis. However, patients undergoing hemodialysis (as identified by hemodialysis events in each database or by dialysis-related procedures) prior to the index surgery were excluded because further progression of their renal dysfunction (i.e., GRF) cannot be determined from their serum creatinine values. Patients were also excluded if they did not have at least one creatinine result from the baseline and postoperative periods, which are described below. Because of this criterion, ambulatory surgical patients that did not have postoperative labs drawn prior to discharge were excluded from the cohort. For patients with multiple surgeries within one hospitalization, only the first surgery was included in the analysis. If patients had multiple separate hospitalizations during the study period, each of these encounters were included. However, once a patient experienced an AKI event, subsequent encounters were excluded.

### Measurement of baseline renal function, AKI and CKD

Baseline renal function was determined by collecting the most recent creatinine value before but not earlier than 6 months before surgery. To determine pAKI, the lowest postoperative creatinine readings included results drawn after the “Anesthesia Stop” event in the anesthetic record and before hospital discharge. Follow-up creatinine results were included from outpatient visits (excluding those obtained during subsequent hospitalizations) from 0-36 months after hospital discharge.

For each creatinine value, glomerular filtration rate (GFR) was calculated from each recorded creatinine using the 2021 CKD-EPI formula to maintain a consistent GFR definition.(17) The Acute Kidney Injury Network (AKIN) classification was used to compare postoperative creatinine measurements to baseline renal function in order to identify patients that experienced pAKI.(18,19) Long term CKD at each endpoint was defined as a decline in follow-up GFR of 40% from baseline, in line with trials of kidney disease progression (15).

### Covariates

Data were also collected for a variety of demographic and comorbidity variables, which were selected based on a review of published risk factors for AKI (1,20–23). These included age, sex, body mass index (BMI), self-reported race, surgical specialty and American Society of Anesthesiologists Physical Status Classification (ASA-PS), history of hypertension, congestive heart failure (CHF), chronic pulmonary disease, diabetes, peripheral vascular disease, and liver disease were also recorded. The comorbidity variables were determined from International Classification of Diseases, Ninth Revision and Tenth Revision codes using the icd package(24) in R version 4.2.3 (R Foundation for Statistical Computing, Vienna, Austria). This package implements coding algorithms published by Quan et al (25).

### Statistical Analysis

Data collection and analysis were performed separately at each site. Patients were grouped based on whether they experienced pAKI. Unadjusted comparisons between the two groups were performed using chi-squared tests for categorical variables and Student’s t-tests or Kruskal Wallis tests for continuous variables based on distributions.

To control for differences in the baseline characteristics of patients in the AKI vs non-AKI cohorts, we performed a propensity score matching analysis. For patients that experienced AKI, we included only the AKI encounter while accounting for the number of surgical encounters prior to the AKI encounter. We fitted a logistic regression propensity model to estimate the probability of having AKI for each individual, using the variables listed above, along with the number of surgical encounters prior to the current encounter. Next, we matched every AKI patient to non-AKI patient with a 1:3 ratio based on the propensity score using the nearest neighbor matching with replacement and a caliper width of 0.1 units. When the balance goal was achieved, we performed Kaplan-Meier analysis to estimate the association of AKI with the time to first 40% decline in GFR, based on the outpatient follow-up laboratory data. Further, a stratified Cox proportional hazards regression model was used to assess the association of AKI with time to first 40% decline in GFR for outpatient follow-up laboratory data while accounting for fluctuations in renal function during the follow-up period. The time periods were stratified into 0-6 months, 6-12 months, 12-24 months, and 24-36 months.

To evaluate the overall effect of AKI across the three sites, meta-analysis of each center’s results was performed. We used the inverse variance method to pool the hazard ratios with 95% confidence intervals of three sites together for each time period, individually. Pooled results are reported as OR with 95% CI using random-effects models where the medical center is considered as fixed effect.

Additionally, a secondary analysis evaluating the interaction between AKI, race, and time was performed. A three-way interaction variable was included in the stratified Cox proportion hazard regression model to assess the association of AKI with time to the first 40% decline in GFR for different race groups during different follow-up time periods. Race was stratified into White or non-White to reduce the complexity of the interaction model, and time was stratified into less than or greater than 6 months.

Given the likelihood of patients not having follow-up lab results within each health system, a sensitivity analysis was performed to assess for factors that may predispose patients to lack of follow-up post-discharge creatine determinations. A weight of inverse probability of having follow up lab data was applied to all analyses. Specifically, we fitted a logistic model to estimate the probability of having follow-up using all baseline variables and their interaction with pAKI. All outcome analyses were then weighed using the inverse of this probability and matching weight.

*A priori* statistical significance was set at p < 0.05. All statistical analysis was performed using R version 4.2.3.

### Secondary endpoints

As a secondary endpoint, each site’s database was also queried for postoperative consults or referrals to the nephrology service. Inpatient and outpatient consults up to 6 months postoperatively were recorded. The rates of nephrology consults were calculated for patients stratified by severity of AKI (as reported by the AKIN score).

## Results

The study included 95,208 patients: 38,061 patients from MSH, 35,725 patients from UM, and 21,422 patients from UCLA. The flowchart of the number of patients included and excluded at each step is shown in Supplemental Figure 1. The number of patients that experienced pAKI were 5211 patients (13.7%) from MSH, 3520 patients (9.9%) from UM, and 2297 patients (10.7%) from UCLA. Full comparison of patient characteristics between the unmatched AKI and non-AKI groups are included in Table 1. At all three sites, the AKI group was older, had a higher proportion of male patients, ASA-PS 4 and 5 patients, lower baseline GFR and higher rates of certain comorbidities, such as CHF, hypertension, diabetes, liver disease, renal disease, and peripheral vascular disease. A higher proportion of patients in the AKI group underwent cardiac, urologic, and transplant surgeries.

**Figure 1.**
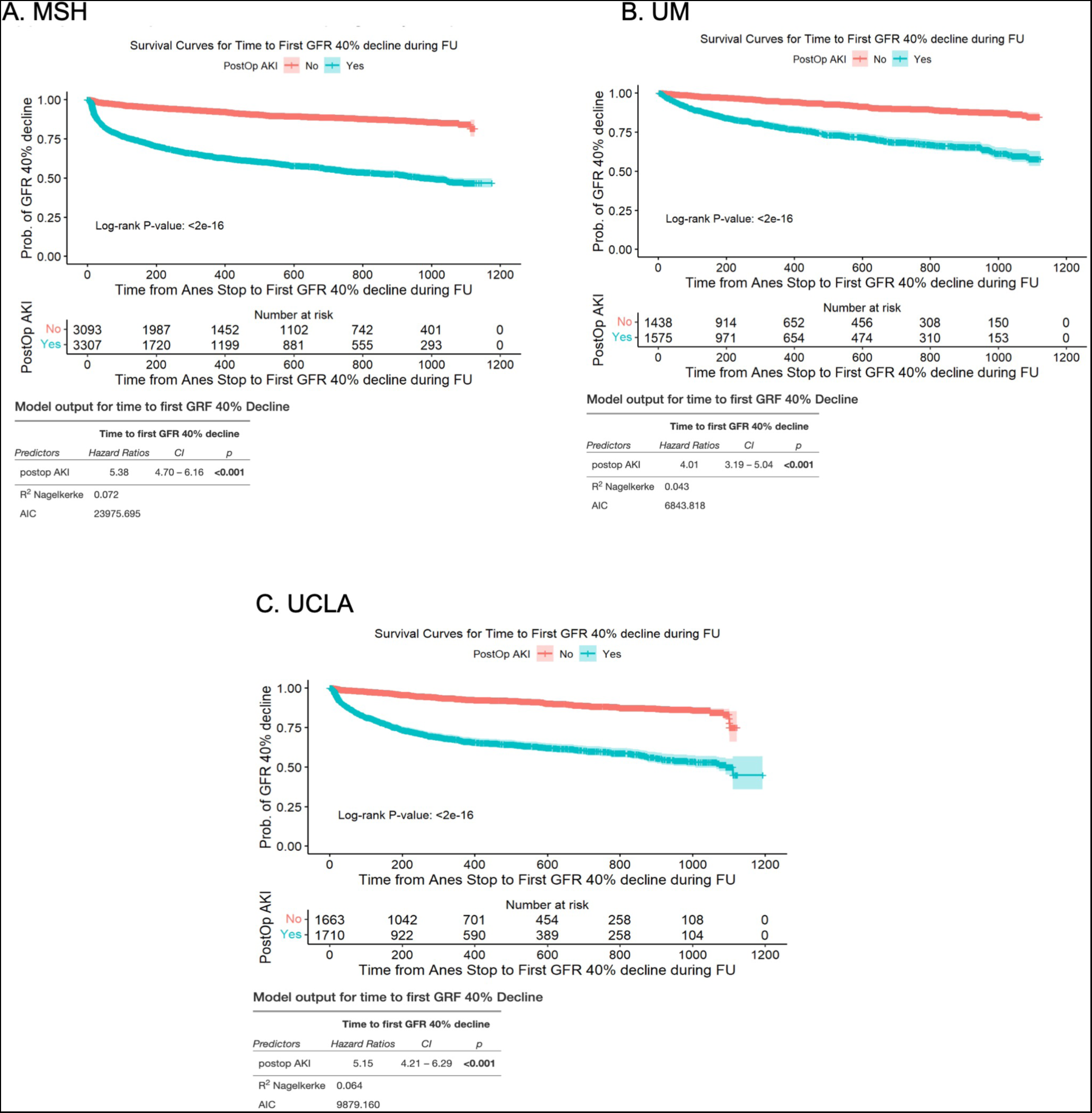
Kaplan-Meier analysis for time to first 40% decline in GFR using propensity score matched sample. MSH: Mount Sinai Hospital, UM: University of Miami, UCLA: University of California Los Angeles

**Table 1.**
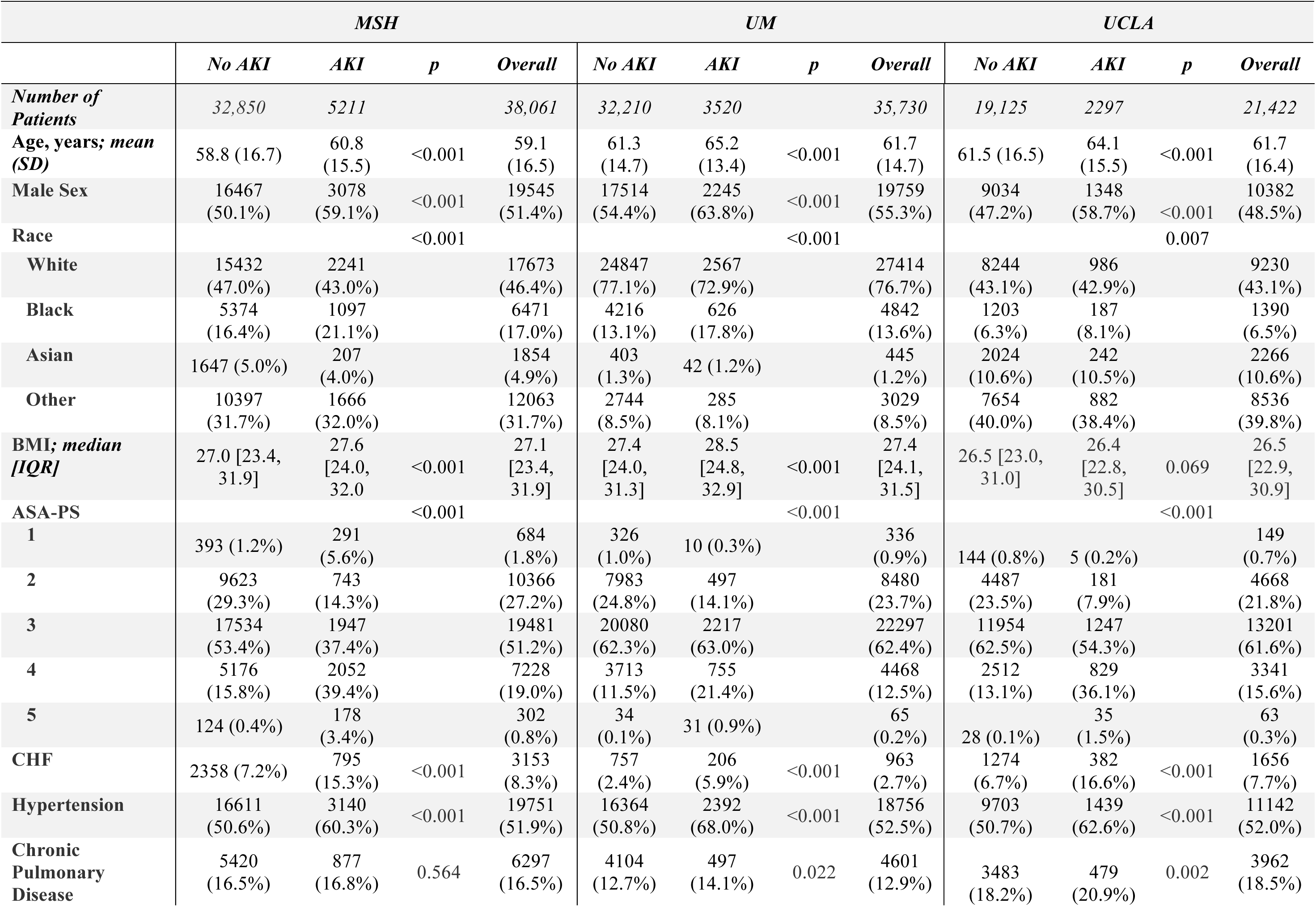
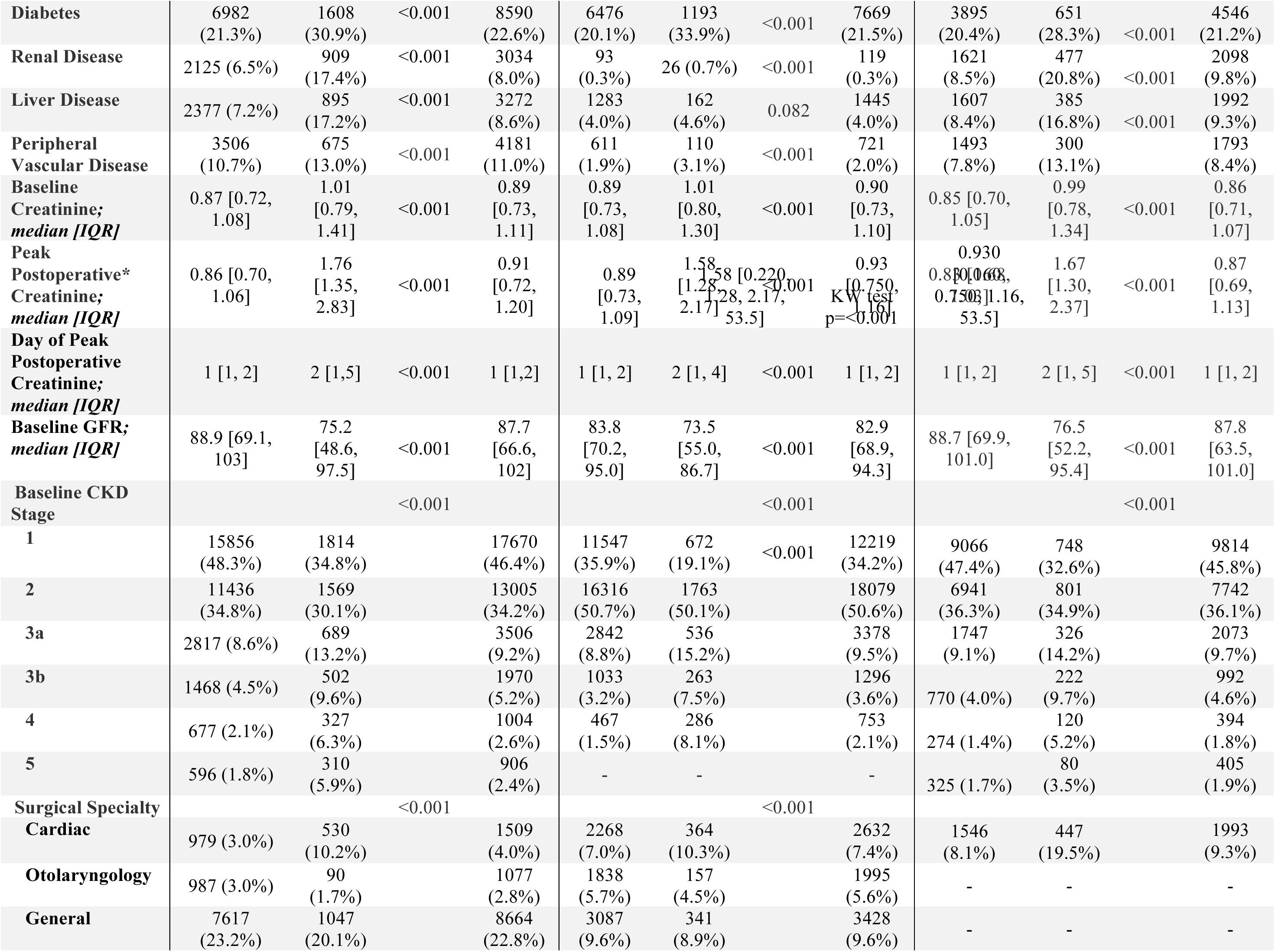
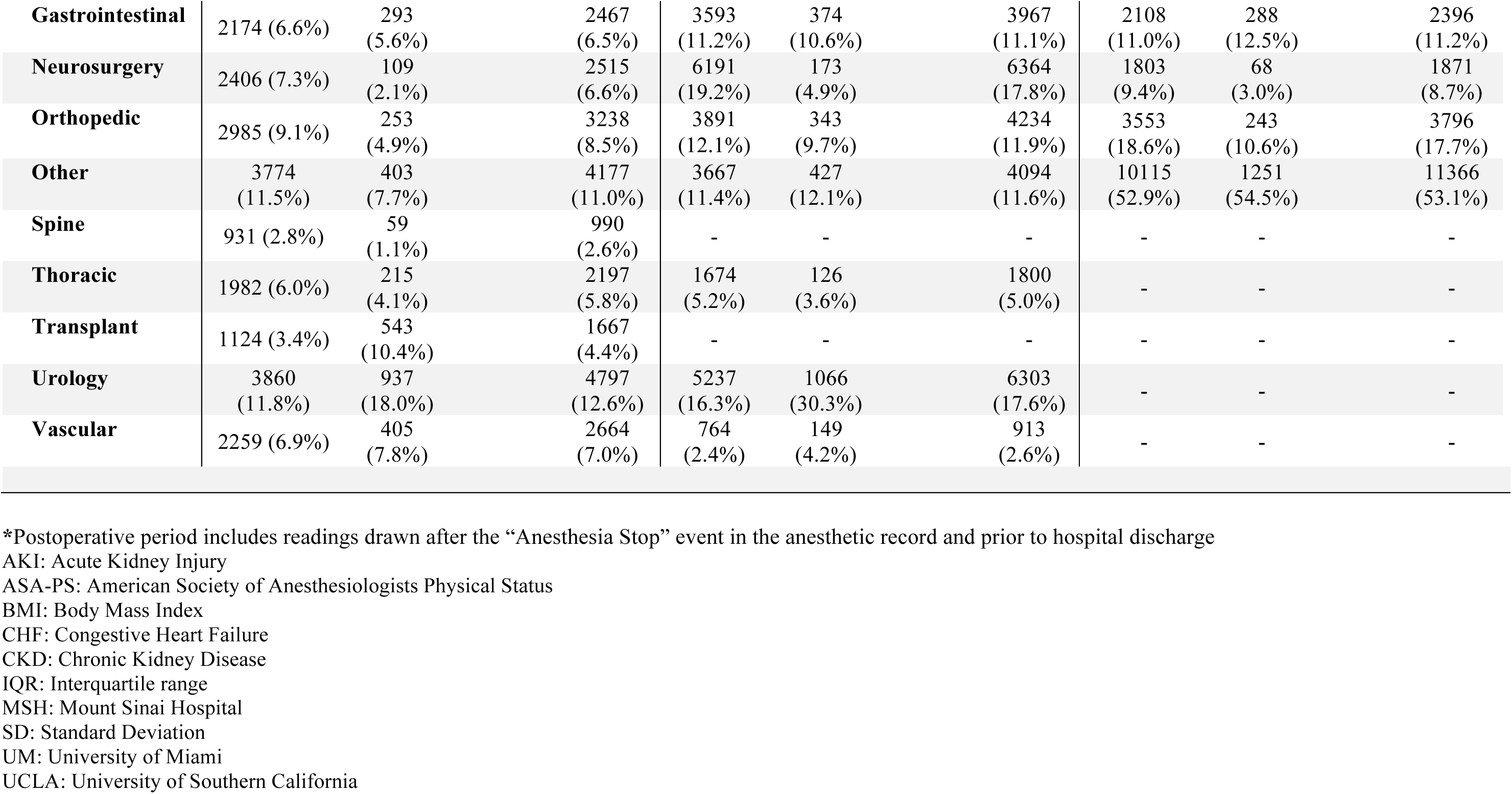
Case Series Characteristics.

|  | *MSH* |  |  |  | *UM* |  |  |  | *UCLA* |  |  |  |
| --- | --- | --- | --- | --- | --- | --- | --- | --- | --- | --- | --- | --- |
|  | *No AKI* | *AKI* | *p* | *Overall* | *No AKI* | *AKI* | *p* | *Overall* | *No AKI* | *AKI* | *p* | *Overall* |
| **Number of Patients** | 32,850 | 5211 |  | 38,061 | 32,210 | 3520 |  | 35,730 | 19,125 | 2297 |  | 21,422 |
| **Age, years; mean (SD)** | 58.8 (16.7) | 60.8 (15.5) | <0.001 | 59.1 (16.5) | 61.3 (14.7) | 65.2 (13.4) | <0.001 | 61.7 (14.7) | 61.5 (16.5) | 64.1 (15.5) | <0.001 | 61.7 (16.4) |
| **Male Sex** | 16467 (50.1%) | 3078 (59.1%) | <0.001 | 19545 (51.4%) | 17514 (54.4%) | 2245 (63.8%) | <0.001 | 19759 (55.3%) | 9034 (47.2%) | 1348 (58.7%) | <0.001 | 10382 (48.5%) |
| **Race** |  |  | <0.001 |  |  |  | <0.001 |  |  |  | 0.007 |  |
| **White** | 15432 (47.0%) | 2241 (43.0%) |  | 17673 (46.4%) | 24847 (77.1%) | 2567 (72.9%) |  | 27414 (76.7%) | 8244 (43.1%) | 986 (42.9%) |  | 9230 (43.1%) |
| **Black** | 5374 (16.4%) | 1097 (21.1%) |  | 6471 (17.0%) | 4216 (13.1%) | 626 (17.8%) |  | 4842 (13.6%) | 1203 (6.3%) | 187 (8.1%) |  | 1390 (6.5%) |
| **Asian** | 1647 (5.0%) | 207 (4.0%) |  | 1854 (4.9%) | 403 (1.3%) | 42 (1.2%) |  | 445 (1.2%) | 2024 (10.6%) | 242 (10.5%) |  | 2266 (10.6%) |
| **Other** | 10397 (31.7%) | 1666 (32.0%) |  | 12063 (31.7%) | 2744 (8.5%) | 285 (8.1%) |  | 3029 (8.5%) | 7654 (40.0%) | 882 (38.4%) |  | 8536 (39.8%) |
| **BMI; median [IQR]** | 27.0 [23.4, 31.9] | 27.6 [24.0, 32.0] | <0.001 | 27.1 [23.4, 31.9] | 27.4 [24.0, 31.3] | 28.5 [24.8, 32.9] | <0.001 | 27.4 [24.1, 31.5] | 26.5 [23.0, 31.0] | 26.4 [22.8, 30.5] | 0.069 | 26.5 [22.9, 30.9] |
| **ASA-PS** |  |  | <0.001 |  |  |  | <0.001 |  |  |  | <0.001 |  |
| **1** | 393 (1.2%) | 291 (5.6%) |  | 684 (1.8%) | 326 (1.0%) | 10 (0.3%) |  | 336 (0.9%) | 144 (0.8%) | 5 (0.2%) |  | 149 (0.7%) |
| **2** | 9623 (29.3%) | 743 (14.3%) |  | 10366 (27.2%) | 7983 (24.8%) | 497 (14.1%) |  | 8480 (23.7%) | 4487 (23.5%) | 181 (7.9%) |  | 4668 (21.8%) |
| **3** | 17534 (53.4%) | 1947 (37.4%) |  | 19481 (51.2%) | 20080 (62.3%) | 2217 (63.0%) |  | 22297 (62.4%) | 11954 (62.5%) | 1247 (54.3%) |  | 13201 (61.6%) |
| **4** | 5176 (15.8%) | 2052 (39.4%) |  | 7228 (19.0%) | 3713 (11.5%) | 755 (21.4%) |  | 4468 (12.5%) | 2512 (13.1%) | 829 (36.1%) |  | 3341 (15.6%) |
| **5** | 124 (0.4%) | 178 (3.4%) |  | 302 (0.8%) | 34 (0.1%) | 31 (0.9%) |  | 65 (0.2%) | 28 (0.1%) | 35 (1.5%) |  | 63 (0.3%) |
| **CHF** | 2358 (7.2%) | 795 (15.3%) | <0.001 | 3153 (8.3%) | 757 (2.4%) | 206 (5.9%) | <0.001 | 963 (2.7%) | 1274 (6.7%) | 382 (16.6%) | <0.001 | 1656 (7.7%) |
| **Hypertension** | 16611 (50.6%) | 3140 (60.3%) | <0.001 | 19751 (51.9%) | 16364 (50.8%) | 2392 (68.0%) | <0.001 | 18756 (52.5%) | 9703 (50.7%) | 1439 (62.6%) | <0.001 | 11142 (52.0%) |
| **Chronic Pulmonary Disease** | 5420 (16.5%) | 877 (16.8%) | 0.564 | 6297 (16.5%) | 4104 (12.7%) | 497 (14.1%) | 0.022 | 4601 (12.9%) | 3483 (18.2%) | 479 (20.9%) | 0.002 | 3962 (18.5%) |
| **Diabetes** | 6982<br>(21.3%) | 1608<br>(30.9%) | <0.001 | 8590<br>(22.6%) | 6476<br>(20.1%) | 1193<br>(33.9%) | <0.001 | 7669<br>(21.5%) | 3895<br>(20.4%) | 651<br>(28.3%) | <0.001 | 4546<br>(21.2%) |
| **Renal Disease** | 2125 (6.5%) | 909<br>(17.4%) | <0.001 | 3034<br>(8.0%) | 93<br>(0.3%) | 26 (0.7%) | <0.001 | 119<br>(0.3%) | 1621<br>(8.5%) | 477<br>(20.8%) | <0.001 | 2098<br>(9.8%) |
| **Liver Disease** | 2377 (7.2%) | 895<br>(17.2%) | <0.001 | 3272<br>(8.6%) | 1283<br>(4.0%) | 162<br>(4.6%) | 0.082 | 1445<br>(4.0%) | 1607<br>(8.4%) | 385<br>(16.8%) | <0.001 | 1992<br>(9.3%) |
| **Peripheral Vascular Disease** | 3506<br>(10.7%) | 675<br>(13.0%) | <0.001 | 4181<br>(11.0%) | 611<br>(1.9%) | 110<br>(3.1%) | <0.001 | 721<br>(2.0%) | 1493<br>(7.8%) | 300<br>(13.1%) |  | 1793<br>(8.4%) |
| **Baseline Creatinine; median [IQR]** | 0.87 [0.72, 1.08] | 1.01<br>[0.79, 1.41] | <0.001 | 0.89<br>[0.73, 1.11] | 0.89<br>[0.73, 1.08] | 1.01<br>[0.80, 1.30] | <0.001 | 0.90<br>[0.73, 1.10] | 0.85 [0.70, 1.05] | 0.99<br>[0.78, 1.34] | <0.001 | 0.86<br>[0.71, 1.07] |
| **Peak Postoperative\* Creatinine; median [IQR]** | 0.86 [0.70, 1.06] | 1.76<br>[1.35, 2.83] | <0.001 | 0.91<br>[0.72, 1.20] | 0.89<br>[0.73, 1.09] | 1.58<br>[1.28, 2.17] | 0.22<br>[0.17, 0.53],<br>KW test<br>p=<0.001 | 0.93<br>[0.75, 1.16] | 0.930<br>[0.80, 1.06],<br>0.750<br>[0.53, 1.16] | 1.67<br>[1.30, 2.37] | <0.001 | 0.87<br>[0.69, 1.13] |
| **Day of Peak Postoperative Creatinine; median [IQR]** | 1 [1, 2] | 2 [1,5] | <0.001 | 1 [1,2] | 1 [1, 2] | 2 [1, 4] | <0.001 | 1 [1, 2] | 1 [1, 2] | 2 [1, 5] | <0.001 | 1 [1, 2] |
| **Baseline GFR; median [IQR]** | 88.9 [69.1, 103] | 75.2<br>[48.6, 97.5] | <0.001 | 87.7<br>[66.6, 102] | 83.8<br>[70.2, 95.0] | 73.5<br>[55.0, 86.7] | <0.001 | 82.9<br>[68.9, 94.3] | 88.7 [69.9, 101.0] | 76.5<br>[52.2, 95.4] | <0.001 | 87.8<br>[63.5, 101.0] |
| **Baseline CKD Stage** |  |  | <0.001 |  |  |  | <0.001 |  |  |  | <0.001 |  |
| **1** | 15856<br>(48.3%) | 1814<br>(34.8%) |  | 17670<br>(46.4%) | 11547<br>(35.9%) | 672<br>(19.1%) | <0.001 | 12219<br>(34.2%) | 9066<br>(47.4%) | 748<br>(32.6%) |  | 9814<br>(45.8%) |
| **2** | 11436<br>(34.8%) | 1569<br>(30.1%) |  | 13005<br>(34.2%) | 16316<br>(50.7%) | 1763<br>(50.1%) |  | 18079<br>(50.6%) | 6941<br>(36.3%) | 801<br>(34.9%) |  | 7742<br>(36.1%) |
| **3a** | 2817 (8.6%) | 689<br>(13.2%) |  | 3506<br>(9.2%) | 2842<br>(8.8%) | 536<br>(15.2%) |  | 3378<br>(9.5%) | 1747<br>(9.1%) | 326<br>(14.2%) |  | 2073<br>(9.7%) |
| **3b** | 1468 (4.5%) | 502<br>(9.6%) |  | 1970<br>(5.2%) | 1033<br>(3.2%) | 263<br>(7.5%) |  | 1296<br>(3.6%) | 770 (4.0%) | 222<br>(9.7%) |  | 992<br>(4.6%) |
| **4** | 677 (2.1%) | 327<br>(6.3%) |  | 1004<br>(2.6%) | 467<br>(1.5%) | 286<br>(8.1%) |  | 753<br>(2.1%) | 274 (1.4%) | 120<br>(5.2%) |  | 394<br>(1.8%) |
| **5** | 596 (1.8%) | 310<br>(5.9%) |  | 906<br>(2.4%) | - | - |  | - | 325 (1.7%) | 80<br>(3.5%) |  | 405<br>(1.9%) |
| **Surgical Specialty** |  |  | <0.001 |  |  |  | <0.001 |  |  |  |  |  |
| **Cardiac** | 979 (3.0%) | 530<br>(10.2%) |  | 1509<br>(4.0%) | 2268<br>(7.0%) | 364<br>(10.3%) |  | 2632<br>(7.4%) | 1546<br>(8.1%) | 447<br>(19.5%) |  | 1993<br>(9.3%) |
| **Otolaryngology** | 987 (3.0%) | 90<br>(1.7%) |  | 1077<br>(2.8%) | 1838<br>(5.7%) | 157<br>(4.5%) |  | 1995<br>(5.6%) | - | - |  | - |
| **General** | 7617<br>(23.2%) | 1047<br>(20.1%) |  | 8664<br>(22.8%) | 3087<br>(9.6%) | 341<br>(8.9%) |  | 3428<br>(9.6%) | - | - |  | - |

|  |  |  |  |  |  |  |  |  |  |
| --- | --- | --- | --- | --- | --- | --- | --- | --- | --- |
| **Gastrointestinal** | 2174 (6.6%) | 293 (5.6%) | 2467 (6.5%) | 3593 (11.2%) | 374 (10.6%) | 3967 (11.1%) | 2108 (11.0%) | 288 (12.5%) | 2396 (11.2%) |
| **Neurosurgery** | 2406 (7.3%) | 109 (2.1%) | 2515 (6.6%) | 6191 (19.2%) | 173 (4.9%) | 6364 (17.8%) | 1803 (9.4%) | 68 (3.0%) | 1871 (8.7%) |
| **Orthopedic** | 2985 (9.1%) | 253 (4.9%) | 3238 (8.5%) | 3891 (12.1%) | 343 (9.7%) | 4234 (11.9%) | 3553 (18.6%) | 243 (10.6%) | 3796 (17.7%) |
| **Other** | 3774 (11.5%) | 403 (7.7%) | 4177 (11.0%) | 3667 (11.4%) | 427 (12.1%) | 4094 (11.6%) | 10115 (52.9%) | 1251 (54.5%) | 11366 (53.1%) |
| **Spine** | 931 (2.8%) | 59 (1.1%) | 990 (2.6%) | - | - | - | - | - | - |
| **Thoracic** | 1982 (6.0%) | 215 (4.1%) | 2197 (5.8%) | 1674 (5.2%) | 126 (3.6%) | 1800 (5.0%) | - | - | - |
| **Transplant** | 1124 (3.4%) | 543 (10.4%) | 1667 (4.4%) | - | - | - | - | - | - |
| **Urology** | 3860 (11.8%) | 937 (18.0%) | 4797 (12.6%) | 5237 (16.3%) | 1066 (30.3%) | 6303 (17.6%) | - | - | - |
| **Vascular** | 2259 (6.9%) | 405 (7.8%) | 2664 (7.0%) | 764 (2.4%) | 149 (4.2%) | 913 (2.6%) | - | - | - |

\*Postoperative period includes readings drawn after the “Anesthesia Stop” event in the anesthetic record and prior to hospital discharge
AKI: Acute Kidney Injury
ASA-PS: American Society of Anesthesiologists Physical Status
BMI: Body Mass Index
CHF: Congestive Heart Failure
CKD: Chronic Kidney Disease
IQR: Interquartile range
MSH: Mount Sinai Hospital
SD: Standard Deviation
UM: University of Miami
UCLA: University of Southern California

### Propensity score matching for AKI

Propensity score matching for pAKI was performed. Supplemental Figure 1 shows the standardized mean differences for included variables before and after matching, and demonstrates a good matching balance (< 0.1 for all variables).

### Association of pAKI with CKD

Table 2 shows the results of the piecewise Cox model for estimating hazard of AKI on developing CKD (defined as a 40% decline in GFR) over a series of postoperative time intervals using the propensity score matched sample. The hazard ratio for the effect of pAKI on 40% decline in GFR was significant throughout the entire 36-month follow-up period at all three sites. In the meta-analysis, the 0-6 month interval had the highest common effect hazard ratio of 13.35 (95% CI: 10.79 to 16.51, p<0.001). The common effect hazard ratio was 7.07 (5.52 to 9.05, p<0.001) for 6-12 months, 6.02 (4.69 to 7.74, p<0.001) for 12-24 months, and 4.32 (2.65 to 7.05, p<0.001) for 24-36 months. The results of the Kaplan-Meier analysis for time to first 40% decline in GFR is shown in Figure 2. The curves show significantly greater time free from 40% decline in GFR for the non-AKI group for all three sites (MSH: HR 5.38, CI 4.70 to 6.16, p<0.001; UM: HR 4.01, CI 3.19 to 5.04, p<0.001; UCLA: HR 5.06, CI 4.21 to 6.29, p<0.001).

**Table 2.** Piecewise Cox model for Estimating Hazard of AKI on Developing 40% Decline in GFR using Propensity Score Matched Sample with Meta-analysis*.

|  |  |  |  |  |
| --- | --- | --- | --- | --- |
|  | Common Effect<br>Hazard Ratio (95% CI) | P value | Site | Site Effect<br>Hazard Ratio (95% CI) |
| **0-6 months** | 13.35 (10.79, 16.51) | <0.001 | Miami | 14.17 (9.53, 21.06) |
|  |  |  | Sinai | 11.06 (7.80, 15.67) |
|  |  |  | UCLA | 15.59 (10.83, 22.46) |
| **6-12 months** | 7.07 (5.52, 9.05) | <0.001 | Miami | 7.12 (3.97, 12.77) |
|  |  |  | Sinai | 6.56 (4.66, 9.23) |
|  |  |  | UCLA | 8.03 (5.09, 12.67) |
| **12-24 months** | 6.02 (4.69, 7.74) | <0.001 | Miami | 4.46 (2.21, 8.98) |
|  |  |  | Sinai | 6.82 (4.77, 9.77) |
|  |  |  | UCLA | 5.69 (3.81, 8.51) |
| **24-36 months** | 4.32 (2.65, 7.05) | <0.001 | Miami | 3.89 (1.63, 9.29) |
|  |  |  | Sinai | 6.05 (4.09, 8.95) |
|  |  |  | UCLA | 4.32 (2.65, 7.05) |

\*Inverse variance meta-analytical method was applied;
AKI: Acute Kidney Injury
CI: Confidence Interval
MSH: Mount Sinai Hospital
UM: University of Miami
UCLA: University of Southern California

### Sensitivity analyses

The percentage of patients that had follow-up outpatient creatinine results in each database was 59% for MSH, 43% for UM, and 72% for UCLA. Because a percentage of patients were lost to follow-up (i.e. did not have serum creatinine values in our database after hospital discharge), a sensitivity analysis was performed using inverse probability weighting for factors that may be associated with patients having available follow-up lab data. After applying this to the model for the effect of pAKI on 40% decline in GFR, similar associations and hazard ratios were observed. The results of this model are shown in Supplemental Table 1.

### Influence of race on CDK after AKI

As reported in Supplemental Table 2, a three-way interaction variable between AKI, race, and time was included in the stratified Cox proportion hazard regression model. The three-way interaction variable was not significant for the MSH (p=0.902) or UM data (0.344) but was significant for the UCLA data (p=0.015). The magnitude of the effect of AKI on risk of GFR decline decreased over time for all racial groups, with more of a decrease in effect for non-White patients than White patients within the UCLA dataset.

### Post-Discharge Nephrology Follow-up

The number of patients with consult notes from the nephrology service up to 6 months postoperatively was recorded, and the rates were stratified by severity of AKI (as reported by the AKIN score). The percentage of patients with postoperative nephrology visits increased with higher AKIN scores. Less than 10% of AKIN 1 and 2 patients had nephrology visits across all three sites. For AKIN 3 patients, the rate ranged from 9.7% at MSH to 21.6% at UCLA. The full rates and times to consult are shown in Supplemental Table 2.

## Discussion

In this study, AKI in the immediate postoperative period was associated renal dysfunction up to 36 months after surgery. This finding was found across three institutions with different procedural and demographic populations. The association was maintained after matching for established risk factors for AKI and also during a sensitivity analysis accounting for missing data. These results demonstrate that even mild pAKI is often not a transient, benign phenomenon, but rather that a clinically significant fraction of such patients do not experience renal recovery back to baseline within several years after the index surgery. This study also showed that fewer than 20% of patients with pAKI saw a nephrologist after discharge, indicating that there may be opportunities for improved follow-up to decrease progression to CKD.

Published rates of AKI vary across published studies, depending both on patient and surgical populations as well as definitions of AKI. The rates of AKI in our study were relatively consistent across the three intuitions studied (13.7% at MSH, 9.9% at UM, and 11.0% at UCLA).

Our study also reaffirmed the demographic, comorbidity, and operative risk factors that have been previously shown to be associated with an increased risk of pAKI (1,20–23).

Similar work focusing on cardiac surgery has demonstrated increased hazard ratios of CKD after pAKI (26). In non-cardiac surgery several studies have shown progression to CKD after pAKI (13,14) however these studies were in smaller single center cohorts. Our study extends this work with a cohort 6 times larger than previously reported, uses a multi-center analysis and risk adjusts for pAKI using propensity score matching. The fact that the progression to CKD remains common after these adjustments indicates that there may be scientific basis to the assumption that pAKI is not only associated with CKD but may actually be an inciting incident in its development.

As part of our analysis, we also assessed the rate of the patients that received a nephrology consult within 6 months postoperatively. The rate of nephrology consults was less than 10% for AKIN 1 and 2 patients and less than 22% for AKIN 3 patients. While the current standard of care is not necessarily to refer patients with pAKI to a nephrologist for follow-up, we believe this may represent an unrecognized opportunity. Just as it is common to refer patients to a cardiologist after an acute cardiac event, referral to a nephrologist may be beneficial in optimizing medication regimens (such as diuretics or anti-hypertensive medications) and ensuring the avoidance of nephrotoxic medications such as NSAIDs which are often common in the postoperative period.

Our study has several limitations. Certain pertinent variables such as postoperative urine output were not reliably available. Some patients did not have follow-up serum creatinine measurements in the institutional EHR database, and the retrospective, deidentified study design made obtaining missing data difficult for patients who may have had followup labs performed elsewhere. However, we performed a sensitivity analysis to assess possible effect of differences between patients with and without documented follow-up of serum creatine and did not find any significant changes in our results. Our study periods were defined to distinguish between inpatient acute changes in renal function and long-term outpatient renal function. However, because only patients with postoperative creatinine results prior to discharge were included, patients undergoing ambulatory surgery that did not have postoperative lab tests drawn before facility discharge were not captured. Thus, our cohort may be skewed toward patients with higher comorbidity burden or undergoing more major surgeries (i.e., requiring inpatient postoperative care) and may also underestimate the true incidence of pAKI. However, the large number of patients and consistent findings across the three institutions support the generalizability of our findings.

In conclusion, this multicenter investigation demonstrated that pAKI was associated with a significantly increased hazard of a persistent 40% decline in GFR up to 36 months after surgery. These findings underline the importance of further study into strategies to help prevent perioperative AKI given its potential for lasting renal dysfunction.

## Data Availability

All data produced in the present study are available upon reasonable request to the authors

