## Supplemental Tables and Figures for "Postoperative Acute Kidney Injury is Associated with Persistent Renal Dysfunction: A Multicenter Propensity Matched Cohort Study"

**Supplemental Figure 1.** Flow chart of inclusion/exclusion criteria

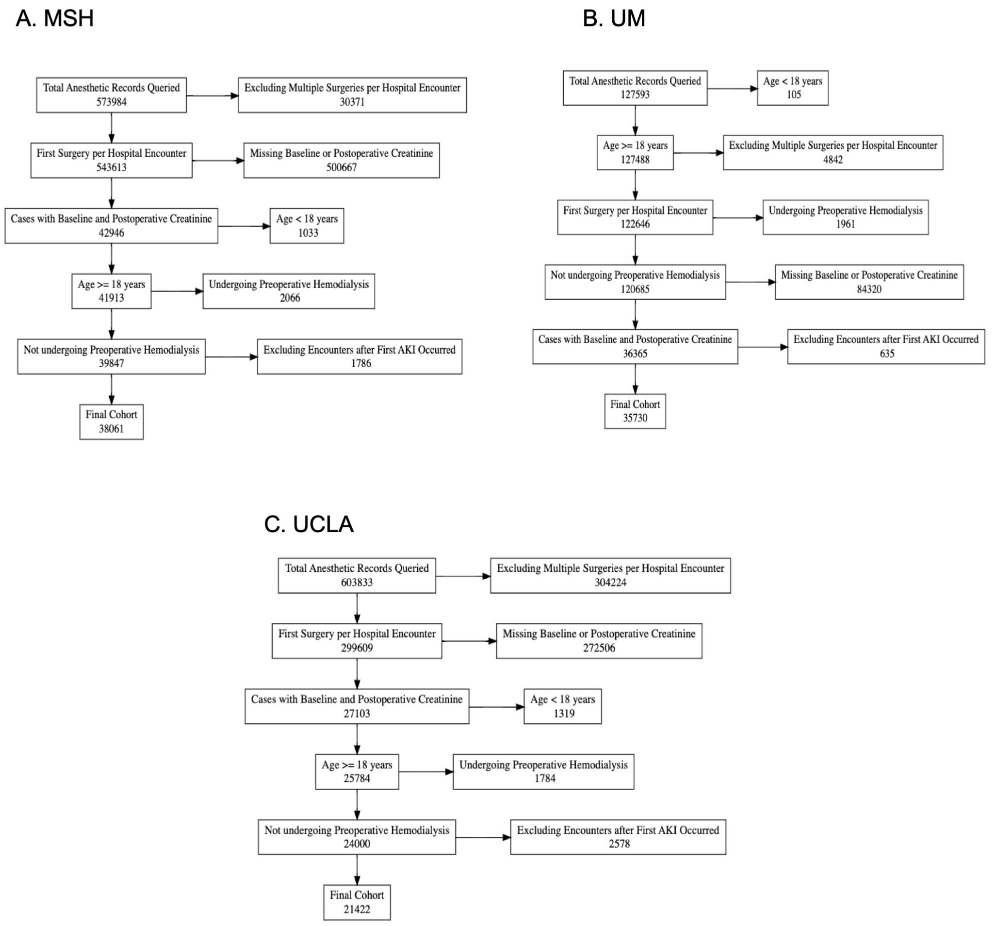

**Supplemental Figure 2:** Standardized mean differences for included variables before and after propensity score matching

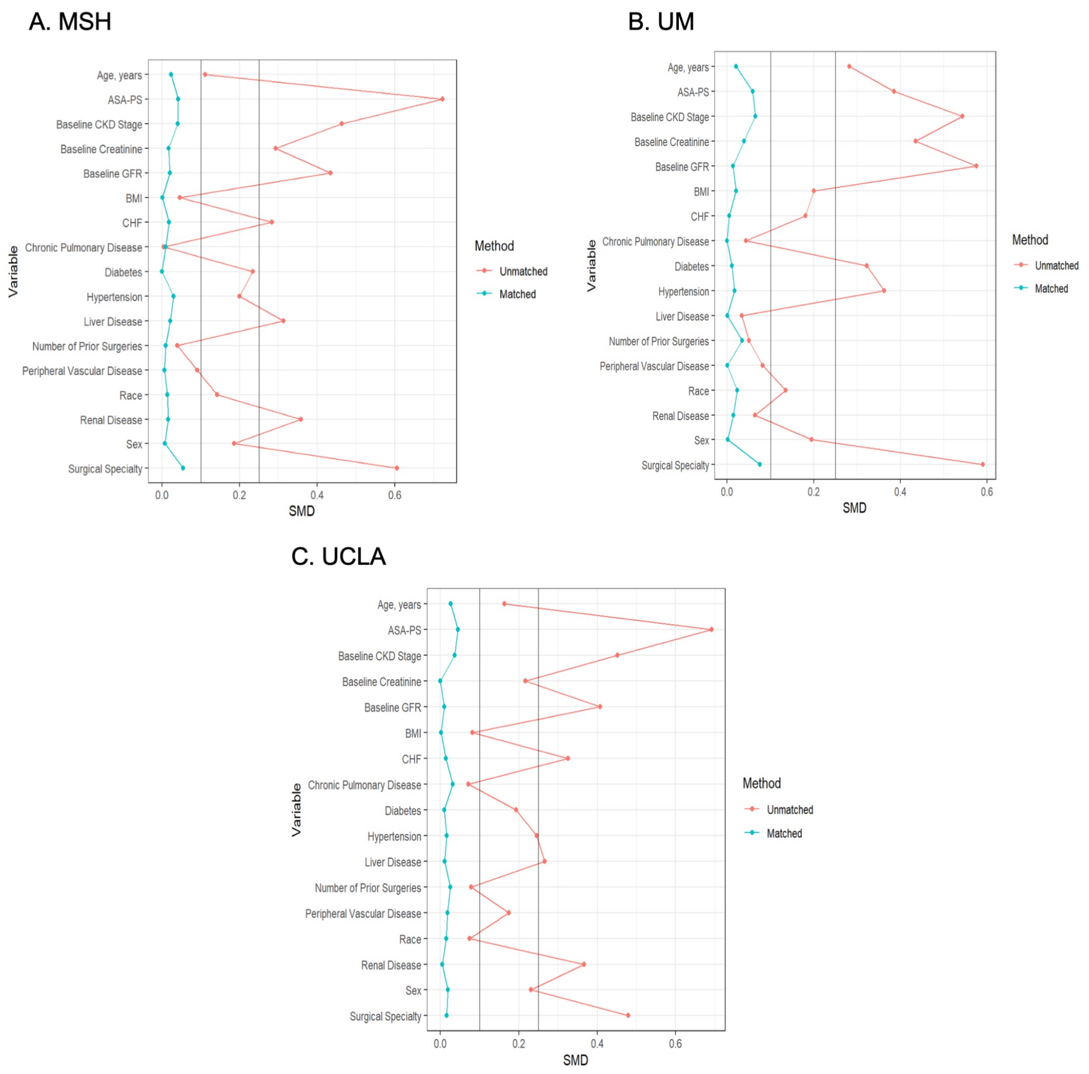

**Supplemental Table 1. Piecewise Cox model for Estimating Hazard of AKI on Developing 40% Decline in GFR using Propensity Score Matched Sample after Applying Weight of Inversed Probability of Having Follow-Up**

| | *MSH* | | | *UM* | | | *UCLA* | | |
|---|---|---|---|---|---|---|---|---|---|
| ***Variable*** | ***Hazard Ratio*** | ***95% Lower CI*** | ***95% Upper CI*** | ***Hazard Ratio*** | ***95% Lower CI*** | ***95% Upper CI*** | ***Hazard Ratio*** | ***95% Lower CI*** | ***95% Upper CI*** |
| **0-6 months** | 11.91 | 8.55 | 16.60 | 12.41 | 8.49 | 18.14 | 15.20 | 10.57 | 21.87 |
| **6-12 months** | 7.23 | 5.13 | 10.19 | 7.53 | 3.31 | 17.15 | 8.07 | 5.10 | 12.76 |
| **12-24 months** | 7.06 | 4.94 | 10.11 | 3.88 | 2.12 | 7.12 | 5.86 | 3.94 | 8.73 |
| **24-36 months** | 6.01 | 4.09 | 8.83 | 3.46 | 1.42 | 8.43 | 4.35 | 2.68 | 7.06 |

AKI: Acute Kidney Injury
CI: Confidence Interval
CKD: Chronic Kidney Disease
GFR: Glomerular Filtration Rate
MSH: Mount Sinai Hospital
UM: University of Miami
UCLA: University of Southern California

**Supplemental Table 2. Piecewise Cox model for Estimating Hazard of AKI on Developing 40% Decline in GFR using Propensity Score Matched Sample after Applying Weight of Inversed Probability of Having Follow-Up**

| | *MSH* | | | *UM* | | | *UCLA* | | |
|---|---|---|---|---|---|---|---|---|---|
| *Variable* | *Estimate* | *Standard Error* | *p* | *Estimate* | *Standard Error* | *p* | *Estimate* | *Standard Error* | *p* |
| **AKI** | 2.61 | 0.18 | **<0.001** | 2.80 | 0.20 | **<0.001** | 2.82 | 0.16 | **<0.001** |
| **Non-White Race** | 0.38 | 0.31 | 0.231 | 0.13 | 0.37 | 0.733 | -0.15 | 0.21 | 0.654 |
| **AKI * Time (> 6 months)** | -0.67 | 0.2 | **0.006** | -0.98 | 0.22 | **0.027** | -0.4 | 0.2 | 0.159 |
| **AKI * Non-White Race** | -0.91 | 0.33 | **0.019** | -0.66 | 0.39 | 0.133 | -0.14 | 0.21 | 0.712 |
| **Time (> 6 months) * Non-White Race** | -0.15 | 0.35 | 0.717 | 0.21 | 0.40 | 0.717 | 0.6 | 0.25 | 0.064 |
| **AKI * Minority * Time > 6 months** | 0.08 | 0.37 | 0.87 | -0.28 | 0.42 | 0.652 | -0.9 | 0.26 | **0.015** |

AKI: Acute Kidney Injury
GFR: Glomerular Filtration Rate
MSH: Mount Sinai Hospital
UM: University of Miami
UCLA: University of Southern California

| Supplemental Table 3. Rates of Postoperative Nephrology Consults Stratified by Severity of AKI | | | | |
|---|---|---|---|---|
| **AKIN Score** | **Site** | **Number of Patients** | **Number of Consults** | **Mean Weeks Until Nephrology Consult Note** |
| **0** | UM | 32,765 | 868 (2.6%) | 10 |
| | MSH | 32,850 | 264 (0.8%) | 7 |
| | UCLA | 21087 | 465 (2.2%) | 10 |
| **1** | UM | 2945 | 278 (9.4%) | 9 |
| | MSH | 3562 | 119 (3.3%) | 6 |
| | UCLA | 1964 | 144 (7.9%) | 9 |
| **2** | UM | 363 | 23 (6.3%) | 8 |
| | MSH | 627 | 32 (5.1%) | 10 |
| | UCLA | 358 | 48 (14.1%) | 8 |
| **3** | UM | 378 | 66 (17.5%) | 7 |
| | MSH | 1022 | 99 (9.7%) | 6 |
| | UCLA | 428 | 90 (21.6%) | 6 |

AKI: Acute Kidney Injury
AKIN: Acute Kidney Injury Network
MSH: Mount Sinai Hospital
UM: University of Miami
UCLA: University of Southern California
